# A comparison of mortality for neonates aged 35 versus 36 weeks gestational age with hypoxic-ischemic encephalopathy treated with therapeutic hypothermia in the National Inpatient Sample

**DOI:** 10.64898/2026.10.19.26365848

**Authors:** Janine Y. Khan, James M. Groh, Keith A. Dookeran

## Abstract

**Objectives:** Using a nationally representative database of U.S. hospitalizations, we aimed to (1) characterize contemporary trends in hypoxic-ischemic encephalopathy (HIE) prevalence by severity, therapeutic hypothermia (TH) utilization, and associated in-hospital mortality; and (2) examine the association between TH and mortality by HIE severity and gestational age (GA), focusing on 35- and 36-weeks GA.

**Methods:** We analyzed newborn hospitalizations from the National Inpatient Sample, 2016-2023. Unspecified, mild, moderate, and severe HIE were identified using ICD-10-CM diagnosis codes P91.60-P91.63; and 35- and 36-weeks GA using codes P07.38-P07.39. TH was identified using ICD-10-PCS codes 6A4Z0ZZ and 6A4Z1ZZ. Survey-weighted multivariable generalized linear models with marginal standardization were used to estimate adjusted mortality risk differences (RD) with surprisal (S) values, and assess whether associations between TH and mortality differed by HIE severity across GA strata.

**Results:** Among 29,767,238 hospital discharges, HIE was identified in 66,535 neonates (2.24 per 1,000), and TH used in 19.2% of HIE cases. In-hospital mortality was 9.74% among all HIE cases and 6.65% among TH-treated neonates. From 2016-23, HIE prevalence nearly doubled (1.58 to 3.01 per 1,000; p-for-trend <0.001) and TH utilization more than doubled (0.33 to 0.74 per 1,000; p-for-trend <0.001), while TH use as a proportion of HIE remained stable (17.8%-20.8%; p-for-trend=0.39). HIE severity distribution was: mild (15%), moderate (29.4%), severe (18.3%), and unspecified (37.3%), with moderate HIE increasing over time (22.3%-33.5%). Crude mortality was consistently highest for severe HIE (range: 33.65%-35.25%). Among infants with severe HIE, TH was associated with reduced mortality overall (RD −10.12%; 95% CI −14.86, −5.39; p<0.001; S=15.12), and specifically at 36-weeks GA (RD −21.98%; 95% CI −35.85, −8.12; p=0.002; S=9.06), but not at 35-weeks (RD −1.83%; 95% CI −21.2, 17.55; p=0.86; S=0.22).

**Conclusions:** TH was associated with mortality reduction in severe HIE overall and at 36-weeks GA, but this benefit remains uncertain at 35-weeks.

## 1 Background

Hypoxic-ischemic encephalopathy (HIE) is an important cause of neonatal mortality and long-term neurodevelopmental impairment following perinatal hypoxia-ischemia. A 2026 American Academy of Pediatrics (AAP) Clinical Report states that therapeutic hypothermia (TH) is standard of care for neonates with moderate-to-severe HIE born at ≥36 weeks gestation, and reduces the risk of death or neurodevelopmental disability^1^. An earlier AAP Report from 2014 acknowledged that regarding inclusion on 6 prior randomized clinical trials (RCTs) of induced hypothermia for neonatal encephalopathy, ‘in all trials infants were at least 35 weeks gestation at birth’, though the total number of 35-week infants across the original trials has been reported to be only seven^2,3^. Clinical practice has drifted toward using TH in infants born before 36 weeks GA, a population in which supporting efficacy and safety data are limited^1,4^, but results from several observational studies support feasibility^5–7^. Faix et al. performed the only prospective RCT evaluating TH in infants born at 33 to 35 weeks GA and found no reduction in death or moderate-severe disability, and raised concern for potential harm^2^. This RCT has been criticized as being inadequately powered to demonstrate benefit (or a lack of benefit) as only 48 infants were enrolled at GA 35 to 35 6/7 weeks^1^. In a recent ‘Key Action Statement’, the AAP stipulates that it does not recommend TH in infants born less than 35 weeks GA; however, limited evidence exists regarding the safety and effectiveness of TH for neonates born at 35 0/7 to 35 6/7 weeks GA^1^.

In the absence of additional data from prospective RCTs, retrospective observational studies of HIE and TH in 35-weeks GA infants have largely focused on mortality, rather than neurodevelopmental outcome, and appear to report conflicting results. El-Dib et al. recently reported that real-world experience from 22 centers shows lower mortality than reported in the Faix et al. RCT specifically for 35-weeks GA, and suggest TH may remain appropriate for select infants of this gestational-age^8^. In contrast to the El-Dib et al. report, Aly et al. recently used Healthcare Cost and Utilization Project (HCUP), 2016–2022, National Inpatient Sample (NIS) data to examine in-hospital mortality in 35-weeks GA infants, and reported that TH use in this subgroup was not associated with differences in in-hospital mortality^9^. This report, however, did not attempt to identify whether TH effects might vary according to HIE severity levels, and appears to show only severity-averaged effects. As such, it remains unknown whether the 35-week mortality estimate reflects a true gestational-age difference in TH efficacy or likely confounding due to variation in HIE severity mix.

National trends in HIE prevalence by severity level and TH utilization have been described through 2018, when mild and moderate HIE diagnoses rose while unspecified HIE declined^10^. Aly et al. described a more recent cohort restricted to 35- and 36-weeks GA infants and reported significant increases in HIE prevalence and TH use through 2022 overall, but did not examine trends by severity level ^9^. No study has updated severity-stratified national trends through the most recent available years, despite unresolved questions about whether rising HIE prevalence reflects true disease increase or evolving diagnostic practice ^11,12^.

We therefore aimed to characterize contemporary, severity-stratified trends in HIE prevalence, TH utilization, and associated in-hospital mortality using national data through 2023, and examine the association between TH and in-hospital mortality within HIE severity strata, with particular focus on whether this association differs between 35- and 36-weeks GA infants.

## 2 Methods

### 2.1 Study Design

This is a retrospective observational study utilizing cross-sectional administrative discharge data from the U.S. HCUP NIS, between 2016 to 2023. NIS is the largest publicly available all-payer U.S. inpatient healthcare database, estimating around 35 million weighted hospitalizations nationally each year. The NIS approximates a 20% stratified sample of all discharges from U.S. community hospitals, and sample size is fixed ^13^. The HCUP NIS database has been previously used to identify and analyze newborn birth hospitalizations as well as maternal delivery hospitalizations, supporting its use for national analyses of neonatal hospitalizations occurring within the maternity-care setting^14^.

### 2.2 Patient Selection and Variable Definitions

The analytic sample was restricted to newborn infant admissions during the first 28 days after birth. International Classification of Diseases, 10th Revision, Clinical Modification (ICD-10-CM) diagnosis, and Procedure Coding System (ICD-10-PCS) procedure codes, were then used to identify specific subpopulations. Infants with unspecified, mild, moderate or severe HIE were identified using codes P91.60, P91.61, P91.62, P91.63, respectively. Use of TH was identified using the procedure codes 6A4Z0ZZ and 6A4Z1ZZ. Infants with GA of 35 and 36 weeks were identified using codes P07.38 and P07.39. Additional study variables included: sex (female/male); insurance payer type (Medicaid and Self-Pay/Private); ecologic median household income quartile based on residents in the patient’s ZIP Code; calendar year; hospital bed size (small/medium/large); and location (rural/urban).

### 2.3 Statistical Analysis

Covariates were found to have <5% missing data and complete case analysis was used. Multivariable generalized linear models with marginal standardization (predictive margins), were used to estimate mortality risk differences (RD) with 95% confidence intervals (CIs), and surprisal [(S), i.e., the p-value expressed in terms of how many consecutive heads (or bit units) would have to be thrown on a fair coin in a single attempt to achieve the same probability] values^15^ associated with TH, for select GA and HIE severity level strata. Final models for predicted mean mortality risk were adjusted for calendar year, sex, insurance payer type, ecologic quartile of the estimated median household income, and hospital bed size, and included an interaction term between HIE and TH covariates. The identity link was pre-specified as the primary model for estimating adjusted RDs, as it directly yields marginal RDs on the probability scale without requiring back-transformation. A logit-link specification was also fit for each model as a robustness check since identity- and logit-link estimates are mathematically equivalent in unadjusted models, but can diverge modestly once covariates are added, as predicted probabilities become link-dependent. Both are reported in Tables 2 and 3, and unless otherwise specified, RDs and S-values cited in the Results section reflect the identity-link estimate.

For models estimating the TH effect within HIE severity level, severity was modeled as a five-level categorical variable [no HIE (reference), mild, moderate, severe, unspecified HIE] in the full-sample analysis, with the average marginal effect for TH estimated separately within each HIE severity level, using the full population (including infants without HIE). In the 35- and 36-week GA strata, mild and moderate severity levels were combined into a single ‘mild/moderate’ category because no deaths were observed among TH-treated infants with mild HIE in either stratum, making a mild-specific within-group TH effect non-estimable; moderate HIE was folded into this combined category for consistency with the grouping used elsewhere (eTables 2–3). Furthermore, in the 35-week stratum, mild/moderate and unspecified severity levels were additionally excluded from the TH-interaction model because no deaths occurred among TH-treated infants in those cells at 35 weeks, restricting the 35-week severity-stratified estimate to the severe stratum only.

All analyses were conducted using Stata statistical software, version 19.5 (StataCorp, College Station, TX). Analyses accounted for the NIS complex survey design, incorporating the hospital-level primary sampling unit (HOSP_NIS), stratum (NIS_STRATUM), and discharge-level weight (DISCWT) variables specified by AHRQ for each data year. Subpopulation-specific estimates (e.g., HIE and TH subgroups) were generated using design-based estimation rather than sample sub-setting, to preserve correct variance estimation. Design-based confidence intervals could not be computed for strata with no observed deaths among TH-treated cases, and these cells were omitted from the corresponding tables. The within-stratum TH-mortality effect was not estimable for these cells, despite TH-treated cases being included in the underlying model, because no deaths were observed among those cases. In accordance with the HCUP Data Use Agreement, discharge counts of fewer than 11 (applying to both unweighted and weighted counts) are required to be suppressed in tables, figures, or text. As such, in eTable 2, mild and moderate HIE severity levels were combined, and calendar years were grouped into four 2-year periods, because several individual year × severity cells fell below the HCUP DUA <11 discharge threshold at finer resolution. In eTable 3, mild and moderate levels were combined for consistency with the severity collapse already necessitated in the multivariable models (see above); this combination also satisfies the DUA threshold. One cell (mild/moderate HIE at 35 weeks GA, eTable 3) remained below the threshold even after combination and is reported as suppressed ("<11").

### 2.4 Ethics Approval

Since HCUP data is deidentified and consistent with HIPAA requirements for use of a limited data set, the University of Wisconsin-Milwaukee Institutional Review Board (IRB) considered this study to be exempt from review.

## 3 Results

### 3.1 Crude Prevalence Trends

During the study period, a total of 29,767,238 neonatal hospital discharges were recorded. HIE was identified in 66,535 neonates, yielding an overall crude prevalence of 2.24 per 1,000 hospitalizations (Table 1), and HIE prevalence was seen to increase over the period (1.58 to 3.01 per 1,000; p-for-trend <0.001). Crude HIE prevalence was observed to be higher for males (2.54 vs. 1.91 for females), Medicaid and Self-Pay insurance payer type (2.32 vs. 2.05 for Private), lowest median household income quartile (2.34 vs. 2.09 for highest) and urban location (2.41 vs. 0.43 for rural). TH was administered to 15,285 neonates, corresponding to an overall crude prevalence of 0.51 per 1,000 hospitalizations, and TH prevalence was also seen to increase over the period (0.33 to 0.74 per 1,000; p-for-trend <0.001). TH was used in 19.2% of HIE cases, and TH use as a proportion of HIE remained stable (17.8%–20.8%; p-for-trend=0.39). Overall, regardless of TH intervention, 6,480 infants with HIE died during hospitalization, resulting in a mortality rate of 9.7%. Crude prevalence of HIE by severity level over time is shown in Figure 1. Overall, HIE severity levels were: mild (15.0%), moderate (29.4%), severe (18.3%), unspecified (37.3%). Over time, the prevalence trend for the severe HIE level remained relatively flat, while the mild HIE level trend increased modestly before stabilizing after 2020. However, the prevalence trend for the moderate HIE increased more substantially over time, while the trend for the unspecified decreased (eTable 1). Crude HIE mortality by severity level over time is shown in Figure 2 (eTable 2) and was substantially worse for those within the severe category, demonstrating a roughly flat trend over time (crude mortality proportion, range: 33.65%–35.25% across the study period). Crude HIE mortality outcomes stratified by severity level and select GA (eTable 3) show that crude mortality remained highest within the severe HIE stratum across GA groups, at 34.59% (95% CI: 27.78, 41.41) among 36-week infants and 33.33% (95% CI: 24.58, 42.09) among 35-week infants.

**Figure 1.**
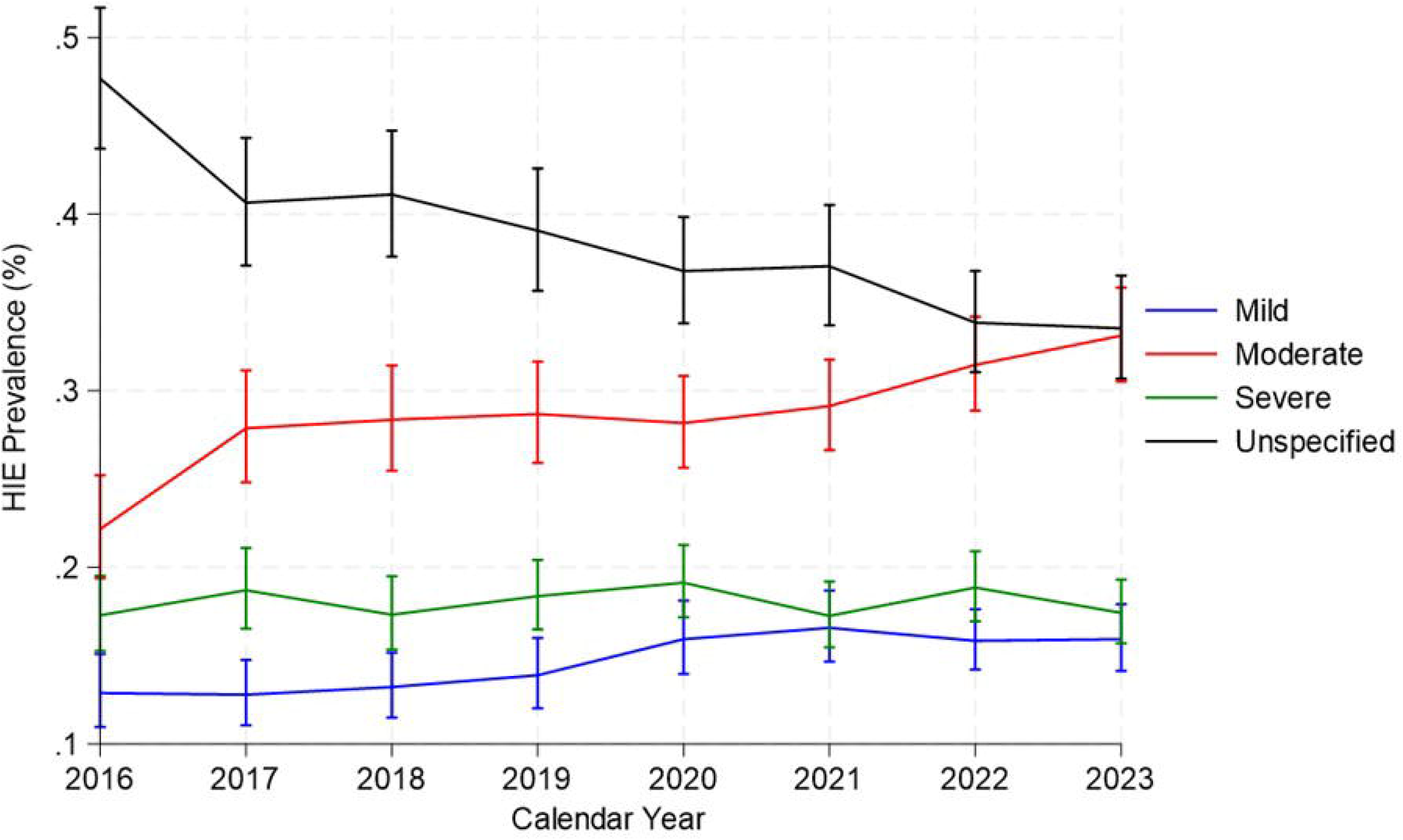
Crude prevalence of HIE by severity level and year in neonates (NIS 2016-23 data).

**Figure 2.**
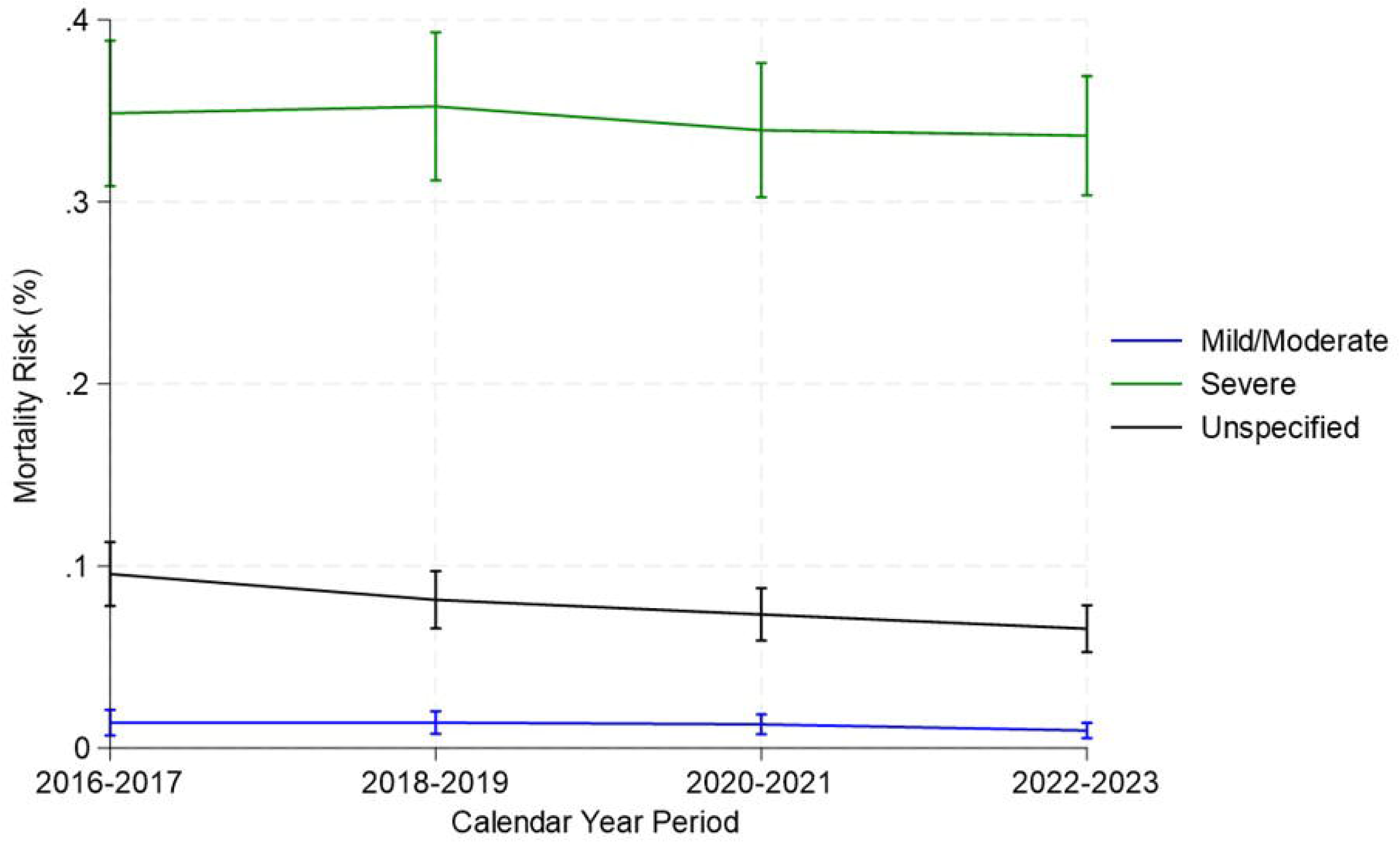
Crude HIE mortality outcomes by severity level and 2-year period (NIS 2016-2023 data).

**Table 1.** Prevalence of HIE and therapeutic hypothermia by select factors in all neonates (NIS 2016-2023 data)

|  |  |  | All neonates<sup>a</sup> | HIE |  |  | Therapeutic Hypothermia |  |  |
| --- | --- | --- | --- | --- | --- | --- | --- | --- | --- |
| FACTORS | (% missing) |  | Weighted Sample Size | Weighted Counts | Weighted Proportions (per 1000 hospitalizations) | 95% CI | Weighted Counts | Weighted Proportions (per 1000 hospitalizations) | 95% CI |
| Year | (0) | 2016 | 3953756 | 6250 | 1.58 | ( 1.42 , 1.77 ) | 1320 | 0.33 | ( 0.27 , 0.42 ) |
|  |  | 2017 | 3863896 | 6925 | 1.79 | ( 1.61 , 2.00 ) | 1480 | 0.38 | ( 0.31 , 0.48 ) |
|  |  | 2018 | 3780399 | 7565 | 2.00 | ( 1.80 , 2.22 ) | 1680 | 0.44 | ( 0.36 , 0.54 ) |
|  |  | 2019 | 3736584 | 7705 | 2.06 | ( 1.86 , 2.29 ) | 1855 | 0.50 | ( 0.41 , 0.61 ) |
|  |  | 2020 | 3587841 | 8130 | 2.27 | ( 2.06 , 2.50 ) | 1960 | 0.55 | ( 0.45 , 0.66 ) |
|  |  | 2021 | 3634342 | 9595 | 2.64 | ( 2.39 , 2.92 ) | 2240 | 0.62 | ( 0.51 , 0.75 ) |
|  |  | 2022 | 3645940 | 9630 | 2.64 | ( 2.40 , 2.91 ) | 2120 | 0.58 | ( 0.48 , 0.71 ) |
|  |  | 2023 | 3564481 | 10735 | 3.01 | ( 2.73 , 3.32 ) | 2630 | 0.74 | ( 0.60 , 0.91 ) |
|  |  | **Total** | **29767238** | **66535** | **2.24** | **( 2.16 , 2.32 )** | **15285** | **0.51** | **( 0.48 , 0.55 )** |
| Sex | (0.06) | Male | 15279264 | 38820 | 2.54 | ( 2.45 , 2.64 ) | 8705 | 0.57 | ( 0.53 , 0.62 ) |
|  |  | Female | 14470339 | 27680 | 1.91 | ( 1.84 , 1.99 ) | 6580 | 0.46 | ( 0.42 , 0.49 ) |
| Payer Type<sup>b</sup> | (3.2) | Private | 13891130 | 28455 | 2.05 | ( 1.96 , 2.14 ) | 7040 | 0.51 | ( 0.47 , 0.55 ) |
|  |  | Medicaid and Self Pay | 14924039 | 34635 | 2.32 | ( 2.23 , 2.42 ) | 7455 | 0.50 | ( 0.46 , 0.54 ) |
| Income Quartile<sup>c</sup> | (0.8) | Quartile 1 (highest) | 6587602 | 13735 | 2.09 | ( 1.97 , 2.21 ) | 3545 | 0.54 | ( 0.48 , 0.61 ) |
|  |  | Quartile 2 | 7345007 | 16175 | 2.20 | ( 2.10 , 2.31 ) | 3830 | 0.52 | ( 0.47 , 0.58 ) |
|  |  | Quartile 3 | 7448687 | 16965 | 2.28 | ( 2.17 , 2.39 ) | 3850 | 0.52 | ( 0.47 , 0.57 ) |
|  |  | Quartile 4 (lowest) | 8148232 | 19045 | 2.34 | ( 2.23 , 2.46 ) | 3910 | 0.48 | ( 0.43 , 0.53 ) |
| Hospital Bed Size | (0) | Small | 5904251 | 10055 | 1.70 | ( 1.55 , 1.87 ) | 1990 | 0.34 | ( 0.28 , 0.41 ) |
|  |  | Medium | 8890117 | 15370 | 1.73 | ( 1.60 , 1.87 ) | 3550 | 0.40 | ( 0.33 , 0.48 ) |
|  |  | Large | 14972870 | 41110 | 2.75 | ( 2.63 , 2.87 ) | 9745 | 0.65 | ( 0.60 , 0.71 ) |
| Died During Hospitalization | (0.02) | No | 29658998 | 60040 | 2.02 | ( 1.95 , 2.10 ) | 14355 | 0.48 | ( 0.45 , 0.52 ) |
|  |  | Yes | 101415 | 6480 | 63.90 | ( 60.34 , 67.65 ) | 925 | 9.12 | ( 7.81 , 10.65 ) |
| Location | (0) | Urban | 27208857 | 65425 | 2.41 | ( 2.32 , 2.49 ) | 15180 | 0.56 | ( 0.52 , 0.60 ) |
|  |  | Rural | 2558381 | 1110 | 0.43 | ( 0.37 , 0.51 ) | 105 | 0.04 | ( 0.02 , 0.07 ) |
|  |  |  |  |  | Weighted Mean |  | Weighted Mean |  |  |
| Length of Stay<sup>d</sup> | (0) | Days |  |  | 16.11 | ( 15.65 , 16.57 ) |  | 16.13 | ( 15.27 , 16.98 ) |

Abbreviations: HIE, hypoxic ischemic encephalopathy; NIS, National Inpatient Sample; CI, Confidence Interval.
<sup>a</sup>All values (totals, count, proportions) are survey weighted.
<sup>b</sup>Excludes other categories.
<sup>c</sup>Ecologic median household income quartile.
<sup>d</sup>Mean length of stay in days for HIE and therapeutic hypothermia groups.

Regarding mortality among neonates with HIE and specifically treated with TH, for all neonates, there were 850 deaths among 12,780 treated (crude mortality proportion 6.65%; 95% CI: 5.67, 7.63); for infants 36 weeks GA there were 75 deaths among 985 treated (7.61%; 95% CI: 3.90, 11.32); and among 35-week GA infants, all observed deaths occurred among those with severe HIE, with no deaths observed among TH-treated infants with mild, moderate, or unspecified HIE, and the small death count in the severe subgroup precludes reporting a crude estimate (eTable 4).

### 3.2 Multivariable Models

Fully adjusted multivariable models (Table 2) demonstrated that TH was associated with lower mortality from HIE in all neonates (RD −4.02%; 95% CI: −5.18, −2.85; p< 0.001; S=35.94). This effect was however less evident for those 36 (RD −2.67%; 95% CI: −7.12, 1.79; p=0.242; S=2.05), and 35 (RD −1.23%; 95% CI: −7.44, 4.99; p=0.697; S=0.52) weeks GA. Fully adjusted multivariable models stratified by HIE severity level (Table 3) suggest that most of the mortality benefit occurs at the severe level; for all neonates (RD −10.12%; 95% CI: −14.86, −5.39; p< 0.001; S=15.12). A smaller mortality benefit was also observed among infants with moderate (RD −1.02%; p=0.006; S=7.39) and unspecified HIE (RD −3.93%; p<0.001; S=14.25), but not mild HIE (RD +0.47%; p=0.363; S=1.46), underscoring that the magnitude of benefit was concentrated in the severe stratum. A mortality benefit at the severe level was also observed at 36 weeks GA (RD −21.98%; 95% CI: −35.85, −8.12; p=0.002; S=9.06). However, at 35 weeks GA, the severity-stratified TH effect could only be estimated for the severe category, and no mortality benefit evident (RD - 1.83%; 95% CI: −21.20, 17.55; p=0.857; S=0.22).

**Table 2.** Multivariable models for the effect of TH on HIE associated mortality in select GA strata (NIS 2016-23 data)

| GA Subgroup | Model | Link<sup>a</sup> | Unweighted N (stratum)<sup>b</sup> | Weighted Population Size (stratum)<sup>b</sup> | AME<sup>c</sup> (TH effect, within HIE=1) | SE | 95% CI | p-value | Surprisal (S = -log<sub>2</sub>(p), bits)<sup>d</sup> |
| --- | --- | --- | --- | --- | --- | --- | --- | --- | --- |
| All | Unadjusted HIE×TH | Identity | 5,952,085 | 29,760,413 | -0.0383 | 0.0058 | ( -0.0496 , -0.0269 ) | <0.001 | 34.37 |
|  |  | Logit (AME) | 5,952,085 | 29,760,413 | -0.0383 | 0.0058 | ( -0.0496 , -0.0269 ) | <0.001 | 34.37 |
|  | Fully adjusted<sup>e</sup> | Identity | 5,712,532 | 28,562,649 | -0.0402 | 0.0060 | ( -0.0518 , -0.0285 ) | <0.001 | 35.94 |
|  |  | Logit (AME) | 5,712,532 | 28,562,649 | -0.0396 | 0.0061 | ( -0.0515 , -0.0277 ) | <0.001 | 33.69 |
| 36 weeks | Unadjusted HIE×TH | Identity | 212,169 | 1,060,845 | -0.0324 | 0.0216 | ( -0.0748 , 0.0100 ) | 0.134 | 2.90 |
|  |  | Logit (AME) | 212,169 | 1,060,845 | -0.0324 | 0.0216 | ( -0.0748 , 0.0100 ) | 0.134 | 2.90 |
|  | Fully adjusted<sup>e</sup> | Identity | 203,600 | 1,018,000 | -0.0267 | 0.0227 | ( -0.0712 , 0.0179 ) | 0.242 | 2.05 |
|  |  | Logit (AME) | 203,600 | 1,018,000 | -0.0257 | 0.0219 | ( -0.0686 , 0.0173 ) | 0.242 | 2.05 |
| 35 weeks | Unadjusted HIE×TH | Identity | 115,625 | 578,125 | -0.0273 | 0.0306 | ( -0.0872 , 0.0327 ) | 0.373 | 1.42 |
|  |  | Logit (AME) | 115,625 | 578,125 | -0.0273 | 0.0306 | ( -0.0872 , 0.0327 ) | 0.373 | 1.42 |
|  | Fully adjusted<sup>e</sup> | Identity | 110,681 | 553,405 | -0.0123 | 0.0317 | ( -0.0744 , 0.0499 ) | 0.697 | 0.52 |
|  |  | Logit (AME)<sup>f</sup> | 110,681 | 553,405 | -0.0076 | 0.0323 | ( -0.0709 , 0.0557 ) | 0.818 | 0.29 |

Abbreviations: HIE, hypoxic ischemic encephalopathy; TH, therapeutic hypothermia; GA, gestational age; NIS, National Inpatient Sample; AME, average marginal effect; SE, standard error; CI, confidence interval.
<sup>a</sup>Specifies the GLM link function used to model the mortality probability; the identity link estimates the risk difference directly from the linear predictor, while the logit link estimates it via the average marginal effect (AME) from the fitted probabilities, so both rows report risk differences on the same scale despite differing in functional form.
<sup>b</sup>Unweighted N and Weighted Population Size reflect the GA stratum's full estimation sample (subpop size for 36- and 35-week strata; full sample for All neonates), not the number of HIE-positive cases; the HIE=1 subgroup is a subset of this population, isolated only in the AME (see note c).
<sup>c</sup>AME - the average marginal effect of TH on mortality probability, expressed as a risk difference, specifically among infants with HIE (i.e., the HIE=1 level from margins, $\text{dydx}(\text{t\_hypo}) \text{ over}(\text{hie})$ ), estimated from a model fit on the full GA-stratum population.
<sup>d</sup>Surprisal $S = -\log_2(p)$ , the Shannon information (bits) carried by the p-value against the null of no TH effect within HIE=1; S is calculated from the exact p-value, which may be substantially smaller than a truncated display value of "<0.001."
<sup>e</sup>Model fully adjusted for sex, payer type, ecologic median household income quartile, year (modeled as a continuous linear term as a categorical specification showed no significant departure from linearity), hospital bed size, and includes HIE\*TH main effects and product term. Both identity- and logit-link AMEs are provided as mathematically identical for the unadjusted HIE×TH model, but likely diverge in the fully adjusted model, since predicted probabilities become link-dependent once additional covariates are included. The fully adjusted sample size is smaller than the unadjusted sample size in each stratum (by approximately 4%) because of listwise deletion on the added covariates; payer type and income quartile account for most of this reduction, missing in approximately 3.2–3.4% and 0.8–0.9% of each stratum, respectively.
<sup>f</sup>The 35-week fully adjusted logit-AME estimate is the least statistically informative in this table ( $S=0.29$ bits) and likely reflects limited power in this stratum rather than a true null effect.

**Table 3.**
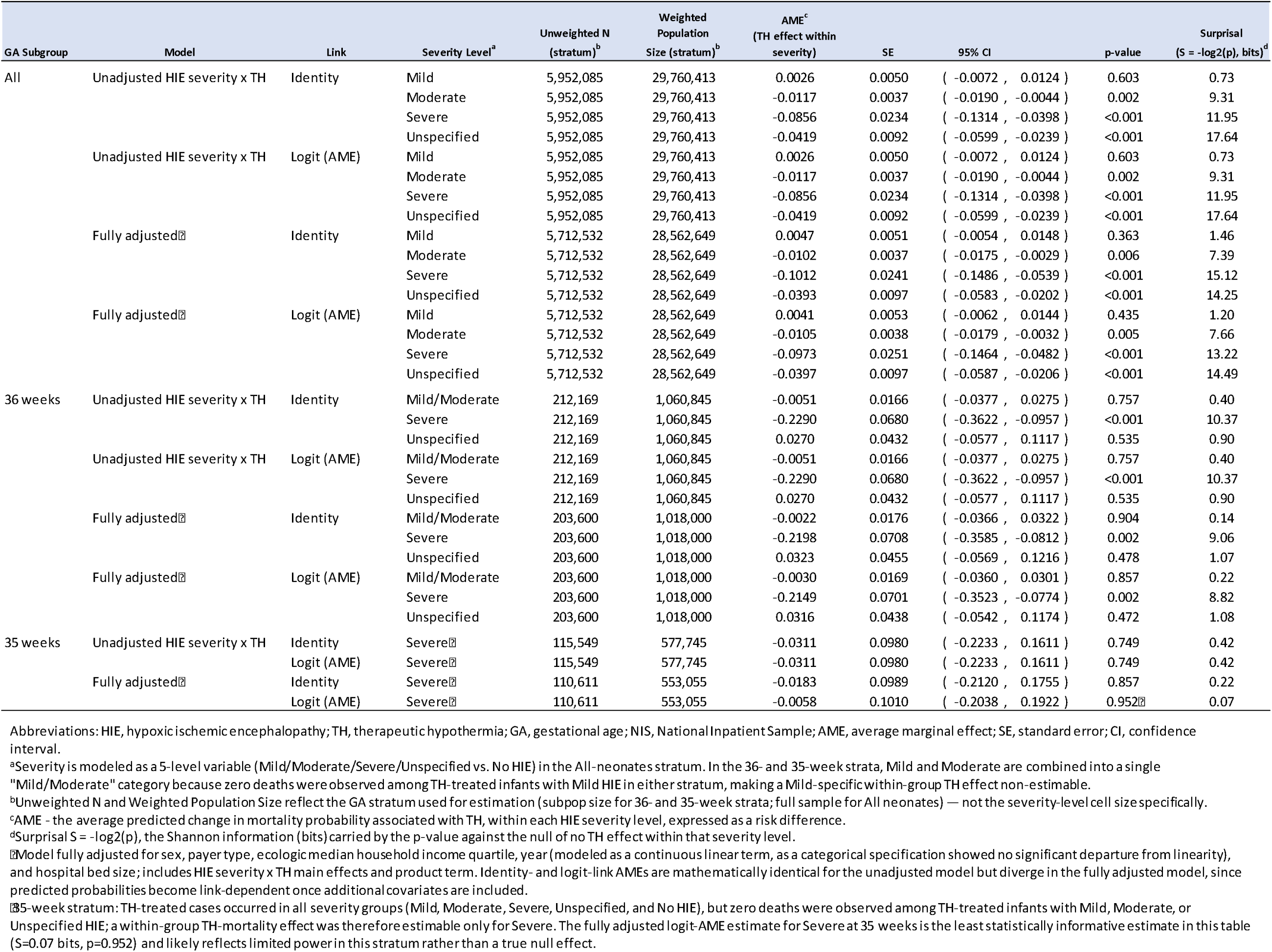
Multivariable models for the effect of TH on HIE associated mortality in select GA strata stratified by severity level (NIS 2016-23 data)

## 4 Comment

### 4.1 Principal findings

In this analysis of more than 29 million neonatal hospitalizations from 2016 to 2023, the crude prevalence of HIE nearly doubled (1.58 to 3.01 per 1,000) and TH use rose in parallel (0.33 to 0.74 per 1,000), while the proportion of HIE cases receiving TH remained stable at approximately 19%. A concurrent shift from unspecified to moderate HIE codes suggests that part of the rise likely reflects improved diagnostic specificity and coding rather than a true change in incidence. Crude in-hospital mortality was concentrated in the severe HIE stratum, affecting roughly one in three infants (33.7%–35.3%) with no improvement across the study period, and was similar at 35- and 36-weeks GA, suggesting that severity rather than GA was the likely dominant determinant of mortality. After adjustment, TH was associated with a 4.0 percentage-point lower mortality risk among all neonates with HIE (S=35.94), but this association was concentrated in the severe HIE stratum (RD −10.1%; S=15.12), with smaller associations in moderate (−1.0%; S=7.39) and unspecified HIE (−3.9%; S=14.25) strata and no evidence of benefit in mild HIE. Among 36-weeks GA infants, the overall association, although suggestive of benefit, was imprecise and compatible with no effect (RD −2.7%; S=2.05); however, the severe stratum showed a larger association (RD −22.0%; S=9.06). At 35 weeks, results were more uncertain, and no association could be established either overall (RD −1.23%; S=0.52) or within the severe HIE stratum (RD −1.8%; S=0.22).

### 4.2 Strengths of the study

Study strengths include use of the largest national population-based database representative of pediatric discharges throughout a diverse range of neonatal units in the U.S. (i.e., HCUP NIS), rather than only select tertiary referral centers. Further, our survey-weighted sample of over 66,000 neonates with HIE over eight years, is a substantially larger cohort than is achievable through single-center or multicenter clinical registries, and one that captures real-world TH utilization patterns. Use of ICD-10-CM/PCS coding also facilitates stratification by HIE severity level and late-preterm GA subgroup.

### 4.3 Limitations of the data

Study limitations, in general, include those inherent to use of administrative discharge data^16^. Specific limitations include the following. First, this is a retrospective analysis of administrative discharge data, and HIE severity level and GA are derived from ICD-10-CM diagnosis codes rather than a validated clinical instrument (e.g., Sarnat staging), introducing the potential for misclassification error and information bias that cannot be directly quantified in this dataset. Second, the unit of analysis was hospital discharges rather than individual newborns. Although a newborn may have more than one admission for the same illness episode, thus introducing a potential source of bias, we consider the likelihood of duplicate admissions is infrequent. We found no duplicate newborns within the analytic dataset. Third, the NIS does not capture longitudinally linked patient-level data (e.g., post-discharge mortality or neurodevelopmental outcomes) and may therefore understate the true mortality burden. Fourth, the NIS contains no information on TH protocol fidelity that could potentially modify treatment effects. Fifth, despite multivariable adjustment for several confounders (i.e., sex, payer type, income quartile, hospital bed size, and year), potential sources of residual and unmeasured confounding likely remain. It should also be noted that covariates included with NIS are subject to recent secular change (for example, race and ethnicity, and census region and division of hospital, have both been removed from the 2023 NIS). Finally, statistical power was limited in the late-preterm subgroups. Although TH-treated infants were present in every severity category at 35 weeks, no deaths occurred among those with mild, moderate, or unspecified HIE, so a within-group TH-mortality association could be estimated only for severe HIE. In addition, for severe HIE, few TH-treated infants were available in these strata, yielding wide confidence intervals and low surprisal values.

### 4.4 Interpretation and implications for practice

The neuroprotective efficacy of TH is established for infants born at or beyond 36 weeks GA through the pivotal RCTs which largely excluded late-preterm infants^17–19^. Evidence for TH at 35 weeks GA is correspondingly limited. Only two earlier RCTs included enrollment at 35 weeks^20,21^, and together they enrolled only seven such infants: two assigned to the non-cooled group (one death, one normal survivor) and five cooled (two deaths, two survivors with moderate disability, and one normal survivor). Although it is plausible that the neuroprotective effect of TH in term and near-term infants extends to late-preterm infants, this assumption remains largely untested.

Observational cohort studies^5,7,22^ suggested that TH was feasible at 34 to 35 weeks GA. This led some centers to offer TH to 35-week infants with moderate-to-severe HIE, resulting in considerable practice variation among neonatologists. These studies were, however, retrospective, small, susceptible to selection bias, and lacked contemporaneous non-cooled control groups, which limits their generalizability. Although their authors concluded that feasibility and short-term complication rates were generally comparable to those of term infants, they also acknowledged that the complications and the combined outcome of death or neurodevelopmental impairment were concerning, warranted cautious use, and required adequately powered RCTs to confirm safety and efficacy.

The only RCT in this population to date, by Faix et al.^2^, randomized 168 infants born at 33 to 35 weeks GA to TH (33.5 °C for 72 hours) or normothermia and found no neuroprotective benefit. Death or moderate-to-severe disability occurred in 35% of cooled and 29% of non-cooled infants (adjusted RR 1.11), and mortality was higher at 20% versus 12% (adjusted RR 1.38). Bayesian analysis estimated a 74% probability of increased death or disability and an 87% probability for death alone with TH. Among a subset of 48 infants born at 35 weeks GA, the results provided no support for TH, but the trial was not powered to estimate a gestation-specific effect. The authors concluded that, until strong supportive evidence emerges from future trials, TH is not indicated for infants with HIE born at ≤35 weeks GA.

El-Dib et al.^8^ offered a counterargument, drawing on the real-world experience of 22 centers in the U.S., Canada, and other regions, and concluded that the cumulative data were reassuring for TH at 35 weeks GA, with no sign of harm and consistent outcomes. Acceptance of these conclusions is limited by the lack of patient-level transparency, potential selection and reporting bias, heterogeneity in follow-up/outcome assessment, and the absence of a normothermic control group. El-Dib et al. also noted baseline imbalances in the Faix RCT, notably a 10-minute Apgar score <5 that was more frequent in the TH group (51% vs 43%). A similar imbalance occurred in the pivotal trials (i.e., TH vs normothermia: CoolCap^18^ 70% vs 55%; NICHD^19^ 80% vs 74%; TOBY^17^ 83% vs 77%), yet infants born at ≥36 weeks GA assigned to TH in those trials consistently had a lower rate of death or disability (CoolCap^18^ 55% vs 66%; NICHD^19^ 49% vs 62%; TOBY^17^ 45% vs 53%), in contrast to the Faix RCT^2^ (i.e., 35% vs 29%). Current practice remains unresolved: in a survey of 54 centers that offer TH to infants born at <36 weeks GA, 33% planned to maintain their protocols, 17% planned to raise GA thresholds or apply stricter eligibility criteria, and 50% were undecided^8^.

Most recently, Aly et al. analyzed in-hospital mortality among 35- and 36-week infants with HIE in the 2016–2022 National Inpatient Sample (NIS)^9^. Neither HIE prevalence (0.48% vs 0.44%) nor mortality (9.98% vs 9.99%) differed between the two GA groups. Among 35-week infants, mortality was 10.3% with TH versus 9.90% without (aOR 0.99; 95% CI 0.69–1.44; p=0.90). Among 35-week infants, moderate HIE was more frequently treated vs. untreated (34.1% vs 25.5%; p=0.12), whereas severe HIE was less frequently treated (7.28% vs. 12.8%; p=0.01). Complications including respiratory distress syndrome, mechanical ventilation >96 hours, pulmonary hemorrhage, pulmonary hypertension, anemia, thrombocytopenia, coagulopathy, and seizures were all more prevalent among TH-treated infants, although length of stay did not differ (median [IQR] 15 [10–23] vs. 13 [3–25] days). The authors concluded that the role of TH at 35 weeks GA remains uncertain and that prospective studies are needed to define safety and eligibility thresholds before routine use can be supported.

Current evidence at 35 weeks GA include a relatively small number of trial participants, a gestation-specific subgroup of one underpowered RCT, retrospective cohorts, uncontrolled real-world data, and one prior national analysis of overall mortality. To our knowledge, this is the first nationally representative study to estimate the absolute mortality difference with TH by both HIE severity and gestational age.

Clinically relevant differences at 35 weeks GA, for example, variation in white matter maturation, cerebral blood flow regulation, and patterns of injury produced by hypoxia-ischemia, may modify the response to cooling^23–25^. In addition, late-preterm infants may be more susceptible to the systemic effects of TH, including impaired thermoregulation, coagulopathy, intraventricular hemorrhage, hypotension and cardiac dysfunction, thrombocytopenia, electrolyte abnormalities, hypo- and hyperglycemia, pulmonary hypertension, and respiratory instability. Neuroprotection demonstrated in infants born at ≥36 weeks GA should therefore not be assumed at 35 weeks, and any benefit may be offset by these systemic risks. Furthermore, recognizing HIE in preterm infants is also inherently difficult. Features used in the modified Sarnat examination, such as hypotonia, feeding difficulty, and apnea, may reflect physiologic immaturity rather than encephalopathy, and amplitude-integrated electroencephalogram (aEEG) background patterns vary with GA and may not represent underlying hypoxic-ischemic change. This diagnostic uncertainty may result in misdiagnosis and therefore introduce misclassification into administrative-code-based definitions of HIE at this GA.

The 2026 AAP clinical report^1^ recommends as standard of care TH for infants born at ≥36 weeks GA with moderate-to-severe HIE, initiated within 6 hours of birth, at a core temperature of 33.5 to 34.5 °C for 72 hours. For infants born at 35 0/7 to 35 6/7 weeks GA, the report advises that cooling be considered case by case after careful discussion with families of the uncertain benefit and potential risks. AAP also states that TH is not recommended below 35 weeks GA outside a research setting. Informed by the Faix et al. RCT^2^ and our findings, TH at 35 weeks GA should not simply be an extension of the term protocol. It should be an individualized, shared decision with parents following thorough counseling about potential treatment harm, and we recommend that informed consent be considered whenever TH is offered at this GA, particularly given the 74% probability of increased death or disability estimated in infants born at 33 to 35 weeks GA^2^. Our findings for severe HIE, overall and at 36 weeks GA, support current guidance for TH in these populations. They also highlight an evidence gap for late-preterm infants <36 weeks who were not included in the pivotal trials, so that target temperatures and safety thresholds for this group are not well established.

The following challenges persist regarding TH use at 35 weeks. First, the evidence base is too sparse to estimate GA- and severity-specific effects at 35 weeks: the only RCT was not powered for gestation-specific analysis, and observational data, including ours, contain few TH-treated 35-week infants with severe HIE. Second, criteria for identifying and grading HIE in late-preterm infants are not well validated, which complicates both eligibility decisions and the interpretation of administrative databases (e.g., NIS). Third, in-hospital mortality represents only one potential outcome; neurodevelopmental outcomes and the morbidity burden of cooling are important modifiable clinical outcome measures that affect subsequent quality of life of survivors of HIE.

### 4.5 Conclusions

Our findings show that TH was associated with in-hospital mortality reduction in severe HIE overall and at 36-weeks GA. In contrast, estimates for 35-weeks infants were indeterminate in every scenario examined, with low S values. In lieu of prospective RCTs, observational retrospective HIE/TH studies in preterm infants have essentially focused on mortality as the primary outcome, however, neurodevelopmental outcome is arguably as important, if not more important, as an outcome measure. Future research in this area should include adequately-powered, prospective randomized controlled trials, or multicenter registries specifically designed to determine feasibility, safety and clinical benefit of improved neurodevelopmental outcome at 35 weeks GA. Furthermore, incorporating validated encephalopathy staging, with modifications of one of the current standardized clinical scoring and staging systems (for example, Sarnat, Modified Sarnat, or Thompson) into prospective research studies would strengthen administrative datasets. Until such evidence is available, the decision to cool a 35-week infant with HIE will necessarily rest on individualized clinical judgment and transparent counseling with families.

## Declarations

### Ethics approval and consent to participate

This study used deidentified admission claims data from a public registry for secondary data analysis (i.e., HCUP), and all data are fully anonymized before access is allowed. Since HCUP data is deidentified and consistent with HIPAA requirements for use of a limited data set, the University of Wisconsin-Milwaukee Institutional Review Board (IRB) considered this study to be exempt from review. Furthermore, our study methodology comports with all Strengthening the Reporting of Observational Studies in Epidemiology (STROBE) guidelines.

### Consent for publication

Not applicable

### Data Availability Statement

The National (Nationwide) Inpatient Sample (NIS) are part of a family of databases and software tools developed for the Healthcare Cost and Utilization Project (HCUP). HCUP inpatient data are based on administrative data—discharge abstracts created by hospitals for billing. The NIS is the largest publicly available all-payer inpatient care database in the United States, containing data on more than seven million hospital stays. The NIS is available for purchase online through the HCUP Central Distributor and subject to HCUP data use agreement and training restrictions. The minimal data sets underlying the results described in this manuscript are from the HCUP Central Distributor and data cannot be shared publicly by the authors due to existing HCUP data use agreement sharing restrictions. Further information on HCUP and data requests, see: https://hcup-us.ahrq.gov/overview.jsp.

### Author Contribution Statement

JYK and KAD made substantial contributions to conception and design of the study. KAD and JMG, made substantial contributions with acquisition and analysis of the study data. All authors made substantial contributions with interpretation of study results. JYK and KAD wrote the first draft of the manuscript. All authors reviewed and approved the final version of the manuscript and agree to be accountable for all aspects of the work.

## Acknowledgments

The study examined inpatient admissions for neonates using discharge data from the 2016-2023 National Inpatient Sample (NIS), HCUP, AHRQ. The results and conclusions are those of the authors and do not necessarily represent the views of AHRQ or the U.S. Department of Health and Human Services Keith Dookeran had full access to all the data in the study and takes responsibility for the integrity of the data and the accuracy of the data analysis.

## Acknowledgment of HCUP Data Partners

Alaska Department of Health

Alaska Hospital and Healthcare Association

Arizona Department of Health Services

Arkansas Department of Health

California Department of Health Care Access and Information

Colorado Hospital Association

Connecticut Hospital Association

Delaware Division of Public Health

District of Columbia Hospital Association

Florida Agency for Health Care Administration

Georgia Hospital Association

University of Hawaii, Hilo Center for Rural Health Science

Hawaii Laulima Data Alliance

Illinois Department of Public Health

Indiana Hospital Association

Iowa Hospital Association

Kansas Hospital Association

Kentucky Cabinet for Health and Family Services

Louisiana Department of Health

Maine Health Data Organization

Maryland Health Services Cost Review Commission

Massachusetts Center for Health Information and Analysis

Michigan Health & Hospital Association

Minnesota Hospital Association (data for Minnesota and North Dakota hospitals)

Mississippi State Department of Health

Missouri Hospital Industry Data Institute

Montana Hospital Association

Nebraska Hospital Association

Nevada Department of Health and Human Services

New Hampshire Department of Health & Human Services

New Jersey Department of Health

New Mexico Department of Health

New York State Department of Health

North Carolina Department of Health and Human Services

North Dakota (data provided by the Minnesota Hospital Association)

Ohio Hospital Association

Oklahoma State Department of Health

Oregon Health Authority

Oregon Association of Hospitals and Health Systems

Pennsylvania Health Care Cost Containment Council

Rhode Island Department of Health

South Carolina Revenue and Fiscal Affairs Office

South Dakota Association of Healthcare Organizations

Tennessee Hospital Association

Texas Department of State Health Services

Utah Department of Health

Vermont Association of Hospitals and Health Systems

Virginia Health Information

Washington State Department of Health

West Virginia Department of Health and Human Resources

Wisconsin Department of Health Services

Wyoming Hospital Association

## Clinical Trial Number

Clinical trial number: not applicable.

## Disclosure of Delegation to Generative AI

The authors declare the use of generative AI in the research and writing process. According to the GAIDeT taxonomy (2026), the following tasks were delegated to GAI tools under full human supervision:

- Proofreading and editing

The GAI tool used was: Claude Sonnet 5.5.

Responsibility for the final manuscript lies entirely with the authors.

GAI tools are not listed as authors and do not bear responsibility for the final outcomes.

Declaration submitted by: Collective responsibility

