## Supplemental Tables for "A comparison of mortality for neonates aged 35 versus 36 weeks gestational age with hypoxic-ischemic encephalopathy treated with therapeutic hypothermia in the National Inpatient Sample" (from another version of this preprint: version 1, 2026-10-21)

### eTable 1. Crude prevalence of HIE by severity level and year in neonates (NIS 2016-23 data)

| | HIE (n= 66535) | Mild | | | Moderate | | | Severe | | | Unspecified | | |
|---|---|---|---|---|---|---|---|---|---|---|---|---|---|
| | Weighted Counts | Weighted Counts | Weighted Proportions | 95% CI | Weighted Counts | Weighted Proportions | 95% CI | Weighted Counts | Weighted Proportions | 95% CI | Weighted Counts | Weighted Proportions | 95% CI |
| **Year** | | | | | | | | | | | | | |
| (% missing - 0) | | | | | | | | | | | | | |
| 2016 | 6250 | 810 | 0.130 | ( 0.110 , 0.152 ) | 1395 | 0.223 | ( 0.195 , 0.254 ) | 1080 | 0.173 | ( 0.153 , 0.195 ) | 2965 | 0.474 | ( 0.434 , 0.515 ) |
| 2017 | 6925 | 890 | 0.129 | ( 0.111 , 0.148 ) | 1945 | 0.281 | ( 0.250 , 0.314 ) | 1300 | 0.188 | ( 0.166 , 0.212 ) | 2790 | 0.403 | ( 0.367 , 0.439 ) |
| 2018 | 7565 | 1005 | 0.133 | ( 0.116 , 0.152 ) | 2170 | 0.287 | ( 0.258 , 0.318 ) | 1330 | 0.176 | ( 0.156 , 0.198 ) | 3060 | 0.405 | ( 0.370 , 0.440 ) |
| 2019 | 7705 | 1070 | 0.139 | ( 0.120 , 0.160 ) | 2225 | 0.289 | ( 0.261 , 0.318 ) | 1450 | 0.188 | ( 0.169 , 0.209 ) | 2960 | 0.384 | ( 0.350 , 0.419 ) |
| 2020 | 8130 | 1300 | 0.160 | ( 0.140 , 0.182 ) | 2335 | 0.287 | ( 0.261 , 0.315 ) | 1575 | 0.194 | ( 0.174 , 0.215 ) | 2920 | 0.359 | ( 0.330 , 0.389 ) |
| 2021 | 9595 | 1615 | 0.168 | ( 0.149 , 0.189 ) | 2855 | 0.298 | ( 0.273 , 0.323 ) | 1700 | 0.177 | ( 0.159 , 0.197 ) | 3425 | 0.357 | ( 0.326 , 0.389 ) |
| 2022 | 9630 | 1535 | 0.159 | ( 0.143 , 0.177 ) | 3050 | 0.317 | ( 0.291 , 0.344 ) | 1835 | 0.191 | ( 0.171 , 0.211 ) | 3210 | 0.333 | ( 0.306 , 0.362 ) |
| 2023 | 10735 | 1730 | 0.161 | ( 0.143 , 0.181 ) | 3600 | 0.335 | ( 0.309 , 0.362 ) | 1900 | 0.177 | ( 0.160 , 0.196 ) | 3505 | 0.327 | ( 0.298 , 0.356 ) |
| ***Overall*** | ***66535*** | ***9955*** | ***0.150*** | ( ***0.143*** , ***0.157*** ) | ***19575*** | ***0.294*** | ( ***0.284*** , ***0.304*** ) | ***12170*** | ***0.183*** | ( ***0.176*** , ***0.190*** ) | ***24835*** | ***0.373*** | ( ***0.362*** , ***0.385*** ) |

Abbreviations: HIE, hypoxic ischemic encephalopathy; NIS, National Inpatient Sample; CI, confidence interval.

Note: All values (Weighted Counts, Weighted Proportions, 95% CI) are survey weighted; estimated using subpop() design-based estimation.

Weighted proportions represent the distribution of HIE severity level within each year's HIE cases.

**eTable 2. Crude HIE mortality outcomes by severity level and 2-year period (NIS 2016-2023 data)[a]**

| Severity Level | Period | Died (Weighted Count)c | Total (Weighted Count) | Weighted Proportion | 95% CI Lower | 95% CI Upper |
|---|---|---|---|---|---|---|
| Mild/Moderate[b] | 2016-2017 | 70 | 5,040 | 1.39% | 0.68% | 2.10% |
| | 2018-2019 | 90 | 6,470 | 1.39% | 0.78% | 2.01% |
| | 2020-2021 | 105 | 8,105 | 1.30% | 0.76% | 1.83% |
| | 2022-2023 | 95 | 9,915 | 0.96% | 0.54% | 1.38% |
| Severe | 2016-2017 | 830 | 2,380 | 34.87% | 30.88% | 38.87% |
| | 2018-2019 | 980 | 2,780 | 35.25% | 31.18% | 39.33% |
| | 2020-2021 | 1,110 | 3,270 | 33.94% | 30.26% | 37.63% |
| | 2022-2023 | 1,255 | 3,730 | 33.65% | 30.37% | 36.92% |
| Unspecified | 2016-2017 | 550 | 5,755 | 9.56% | 7.80% | 11.31% |
| | 2018-2019 | 490 | 6,020 | 8.14% | 6.56% | 9.72% |
| | 2020-2021 | 465 | 6,340 | 7.33% | 5.89% | 8.77% |
| | 2022-2023 | 440 | 6,715 | 6.55% | 5.26% | 7.84% |

Abbreviations: HIE, hypoxic ischemic encephalopathy; NIS, National Inpatient Sample; CI, confidence interval.

[a]All values (Died, Total, proportions) are survey weighted; estimated using subpop() design-based estimation.

[b]Mild and Moderate HIE severity levels are combined here, and years are grouped into 2-year periods, because several individual year x severity cells fell below the HCUP Data Use Agreement <11 discharge suppression threshold at finer resolution (i.e., annual breakout, or Mild/Moderate reported separately). Every cell in this table meets the HCUP DUA requirement that reported counts (i.e., unweighted/weighted counts be >/=11).

[c]Excludes 3 unweighted HIE cases (2 Severe, 1 Unspecified; ~15 weighted) with missing vital status at discharge; these cases are otherwise included in eTable 1's prevalence counts.

**eTable 3. Crude HIE mortality outcomes stratified by severity level and select GA in neonates (NIS 2016-2023 data)[a]**

| Severity Level | GA Subgroup | Died (Weighted Count) | Total (Weighted Count) | Weighted Proportion | 95% CI Lower | 95% CI Upper |
|---|---|---|---|---|---|---|
| Mild/Moderate[b] | All | 360 | 29,530 | 1.22% | 0.95% | 1.49% |
| Severe | | 4,175 | 12160[c] | 34.33% | 32.47% | 36.19% |
| Unspecified | | 1,945 | 24830[c] | 7.83% | 7.08% | 8.59% |
| Mild/Moderate | 36 weeks | 55 | 2,255 | 2.44% | 1.02% | 3.86% |
| Severe | | 320 | 925 | 34.59% | 27.78% | 41.41% |
| Unspecified | | 120 | 1,675 | 7.16% | 4.42% | 9.91% |
| Mild/Moderate | 35 weeks | <11* | 1,140 | suppressed* | suppressed* | suppressed* |
| Severe | | 185 | 555 | 33.33% | 24.58% | 42.09% |
| Unspecified | | 115 | 1,220 | 9.43% | 5.77% | 13.08% |

Abbreviations: HIE, hypoxic ischemic encephalopathy; NIS, National Inpatient Sample; CI, confidence interval; GA, gestational age.

aAll values (Died, Total, proportions) are survey weighted; estimated using subpop() design-based estimation.

[b]Mild and Moderate HIE severity levels are combined, consistent with the collapse already used for the 35-/36-week severity-stratified multivariable models (Table 3).

[c]Excludes 3 unweighted HIE cases (2 Severe, 1 Unspecified; ~15 weighted) with missing vital status at discharge; these cases are otherwise included in eTable 1's prevalence counts.

*Mild/Moderate, 35 weeks: unweighted death count <11, which remains below the HCUP Data Use Agreement suppression threshold even after combining Mild and Moderate. The Died count is masked per the HCUP DUA; Weighted Proportion and 95% CI are also suppressed.

**eTable 4. Crude mortality outcomes for select GA subgroups among neonates with HIE and therapeutic hypothermia (NIS 2016-2023 data)[a]**

| GA Subgroup[b] | Died (Weighted Count) | Total (Weighted Count) | Weighted Proportion | 95% CI Lower | 95% CI Upper |
|---|---|---|---|---|---|
| All neonates | 850 | 12,780 | 6.65% | 5.67% | 7.63% |
| 36 weeks | 75 | 985 | 7.61% | 3.90% | 11.32% |
| 35 weeks | <11* | 525 | suppressed* | suppressed* | suppressed* |

Abbreviations: HIE, hypoxic ischemic encephalopathy; TH, therapeutic hypothermia; NIS, National Inpatient Sample; CI, confidence interval; GA, gestational age.

[a]All values are survey weighted; estimated using subpop() design-based estimation

[b]GA is derived from ICD-10-CM data.

*35 weeks: unweighted death count <11, below the HCUP Data Use Agreement suppression threshold. The Died count is masked per the HCUP DUA; Weighted Proportion and 95% CI are also suppressed. All observed deaths in this stratum occurred among infants with severe HIE; zero deaths were observed among TH-treated infants with mild, moderate, or unspecified HIE at 35 weeks.
